# The impact of mobility restriction measures on the reproduction index of Covid-19 in the city of Queretaro, Mexico

**DOI:** 10.1101/2020.09.06.20189373

**Authors:** Oscar San Roman Orozco, Santiago Agraz Orozco, Isidro A Gutierrez Alvarez, Vasiliki Radaios

**Author notes:** Corresponding Author: 4036 Miller Drive, Glenview Illinois 60026.

## Abstract

An observational study based on official data (CONACYT and Ministry of Health) was carried out in which the effective reproduction index R(e) and the reproduction index R0 are compared with the mobility presented by Google. Additionally, an overview of the development of the pandemic in Querétaro, Mexico. Highlights key events; such as the main government interventions and social factors that could affect society’s behavior.

A positive relationship is observed between Re, R0, and the levels of mobility presented by Google. This indicates that an increase in mobility is associated with the transmission of SARSCoV-2. In February, a significant decrease in mobility is observed, which lasts until approximately May 1st. This period corresponds to an R0 and R(e) between 1.17 and 1.87. After May 1st, there is a sustained increase in mobility levels. And, as of May 16, the effective reproduction index R (e) and the reproduction index R0 begin to increase. This is expected as it reflects the delay between the infection and the diagnosis of COVID-19. The R0 and R (e) increase from 1.45 on May 16 to 3.59 on July 5. According to the baseline of normal mobility levels, an increase from −49.6% on May 1st, to −20.6% on July 5 was observed.

Based on these data, we conclude that the relaxation of restrictive mobility measures should be reconsidered. Despite this, mobility restrictions must not be a unique mitigation strategy for controlling the Reproductive Index. A comprehensive approach is needed, which generates socio-behavioral changes that allow a further reduction in reproductive rates.

**Key Messages:**

- There is a positive correlation between the reproductive number (R0) and the effective reproductive index (Re) of SARS-CoV-2 and mobility trends in Querétaro, Mexico.
- A clear increase in mobility in the population in Queretaro, Mexico is associated with an increase in the rate of transmission of Covid-19.

---

The new coronavirus (SARS-CoV-2) that emerged from Wuhan, China, in December 2019 rapidly spread through Hubei province and the rest of China, now spreading to more than 200 countries with serious repercussions across all social spheres^7^.

The first case in Mexico was reported on February 28, 2020 in Mexico City, carried by a 35year-old man arriving from Italy^5^. Days later, on March 6, the first case of Covid-19 was recorded in Querétaro, imported from a 43-year-old man from Spain^6^. The World Health Organization subsequently declared the situation a pandemic on March 11, 2020.

When faced with this situation, public health research and action became extremely important. The main objective is the prevention of the spread of this disease and interrupting its territorial expansion using tools such as isolation and quarantine, social distancing, and community containment^1,3^. This is accomplished using the knowledge of the natural history of the disease, as well as its mechanisms of person-to-person transmission recognized by various reports that have been written since the onset of the crisis in affected countries.

Covid-19 is caused by the SARS-CoV-2 virus, a beta-type coronavirus. The average incubation period varies from 2–14 days with an average duration of 5.6 days^4^. It is estimated that the infectious period begins 2.3 days before the onset of symptoms, with a peak at 7 days and a decline in the following 7. For the 15th day after the onset of symptoms, 30% of patients are negative for SARS-Cov-2 by PCR, 68% for day 21, 88% for day 28 and 95% for day 33.^4^

Person-to-person transmission can be measured by the basic reproduction index (R0), or in a broader sense the effective reproduction number (Re) that models how many infections can be derived from a case. The SARS-CoV-2 virus has a reproduction rate of 2.6 with a range of (2.1 to 5.1).^8^

This factor depends on and is influenced by the movement of people, as the virus is transmitted by respiratory droplets when an infected person coughs, sneezes or simply speaks. Because the drops normally fall less than 2 meters away, the transmission risk is reduced by maintaining that distance. An important factor to be taken into account in the transmission of the virus is its persistence on surfaces since SARS-Cov-2 can remain on plastic, cardboard and steel for up to 15 days^15^. Furthermore, another important factor for the transmission of this virus is the contagion by contact with an asymptomatic or pre-symptomatic case, evidence that 40 to 50% of cases may be attributable to contact with asymptomatic or pre-symptomatic cases^1,10^.

Different studies show a relationship in disease reproduction rate and mobility in a given region^9,11,19,20^. It is estimated that a 10% reduction in mobility in a region can decrease the reproduction rate from 0.04 to 0.09 points. Since there is no drug treatment for Covid-19, governments around the world have chosen to implement non-pharmacological measures to limit mobility in order to prevent excessive growth of contagion and consequently, the saturation of health systems^9^.

## Methodology

This report describes the epidemic in the state of Querétaro, Mexico using three epidemiological measures: the new daily cases, the basic reproduction rate (R0) and the effective reproduction number (Re). We analyze the relationship between transmission rates, using the reproductive index, and mobility levels using Google’s “Local Mobility Reports”.

The model used for data logging and analysis is a SIR (Susceptible, Infected, Recovered) compartment model described by Kermack and McKendrick^13^. The objective for epidemic analysis is to describe those susceptible to infection, those infected and those recovered^11^. The basic rate of reproduction (R0) indicates the average number of new infections caused by an infected individual during the course of the disease; it is the relationship between the effective contact rate and the rate of recovery^11^. A decrease in the effective contact rate as a result of government and public health strategies decreases R0 and flattens the contagion curve^14^. Similarly, the effective reproduction number was calculated by multiplying R0 by the proportion of susceptible over the population total (Re x R0 x (S/N S I R). This in order to display that as the proportion of susceptible individuals decreases (S/N), transmission is discouraged. These indicators were used because the evidence indicates they are preferred to describe the pandemic^9,19,20^.

The data used was based on the Federal Government’s “COVID-19 Tablero México” with data from the General Directorate of Epidemiology (Dirección General de Epidemiología)^16^. Mobility data was obtained from Google’s “Local Mobility Reports”. Google collected the data anonymously, creating aggregated user data reports that have activated location history and categorized mobility into 6 categories (stores and leisure, supermarkets, pharmacies, parks, transportation stations, workplaces, and residential areas) where visits to places with similar characteristics are grouped together on each day of the week. To obtain a benchmark, a baseline is determined by the average mobility value measured during the period 5 weeks from January 3 to February 6, 2020 (the period prior to the arrival of SARS-CoV-2 in Querétaro). For this model, 5 variables of mobility data were taken (shops and leisure, supermarkets and pharmacies, parks, transportation stations, and workplaces). We exclude residential mobility as residential values are different to the other variables.

This comparison helps us verify the effect of government-implemented mobility restrictions against mobility and the impact on the R0/Re of the virus. Using official and validated mobility and infection data to analyse changes in reproductive rates is a great and relevant parametric because of the high degree of confidence of controlling the pandemic in the state^17^.

## Observations on Association Between Mobility Levels & SARS-CoV-2 Reproductive Index

Based on the data, we observed a strong correlation between increases in mobility levels and subsequent increases in transmission rates of SARS-CoV-2.

Figure 1 depicts the number of cases, recovered, and deaths in Querétaro, Mexico, from the beginning of the epidemic in Mexico in March 5 until July 5, alongside the dates of important events and measures that were taken. It is important to note that social distancing measures were first suggested on March 14, and then became mandatory on March 23, while the use of face masks became mandatory on April 2^13^. The number of daily new cases did not exceed 15. Between May 5 and May 10, we observed a sharp incline in the number of cases, from 10 daily cases to 47 cases. From May up until July 5 the daily number of cases in Querétaro remain relatively high, especially when compared to the number of cases in April when there were strict restrictions on mobility.

**Figure 1:**
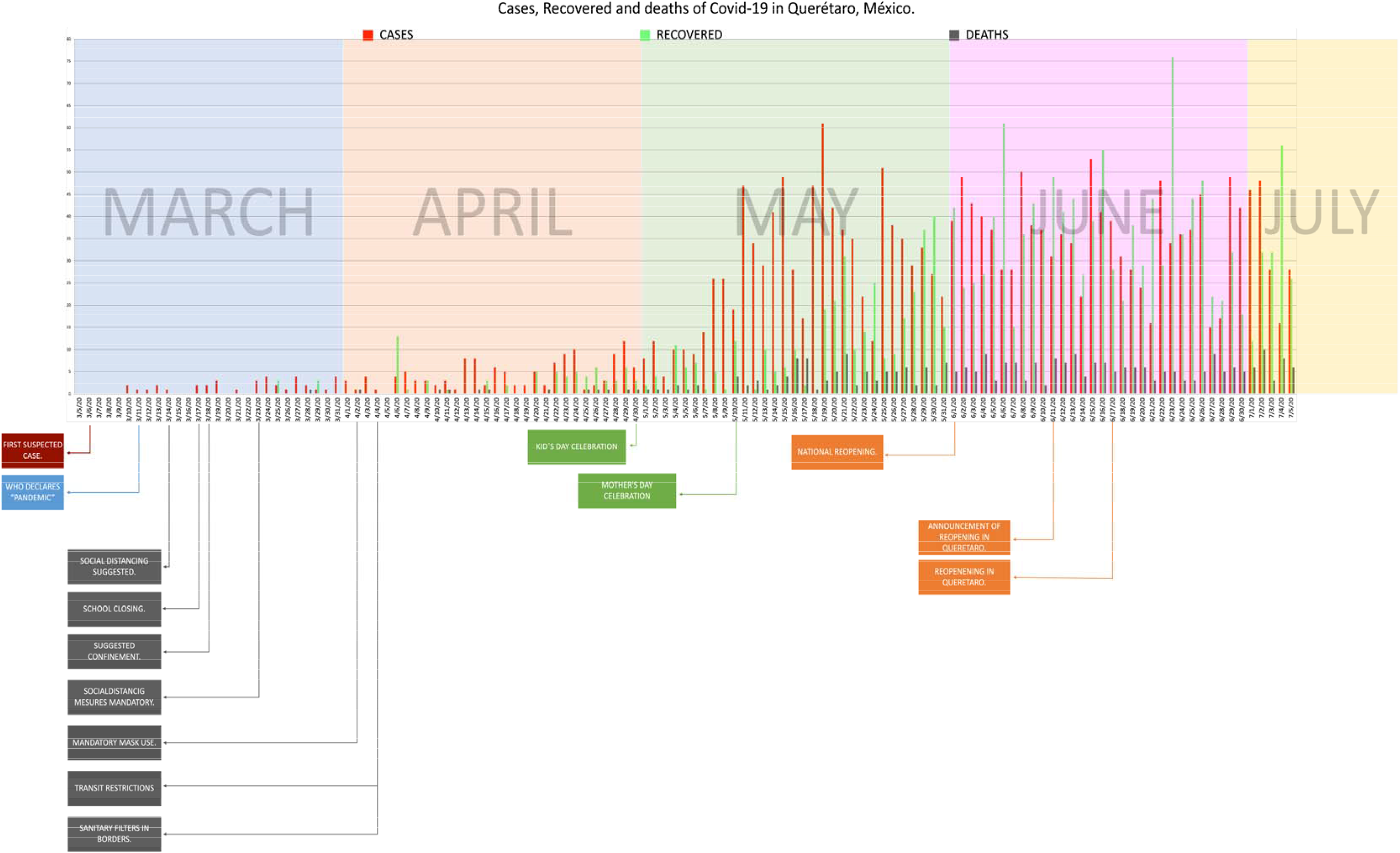
History of SARS-CoV-2 in Querétaro México

Figure 2 and 3 depict the trends in mobility using unidentifiable data from Google. The information on mobility levels during the time period of late February up until July 5 is separated into five categories: recreation, supermarkets and pharmacies, parks, transportation stations, and workplace. Figure 3, depicting average mobility levels indicates a clear and sharp decrease in mobility in Querétaro’s population, with the lowest mobility on April 9. Long before it was announced that activities returned in Querétaro on June 11, mobility began to steadily increase from the beginning of May and into July.

**Figure 2:**
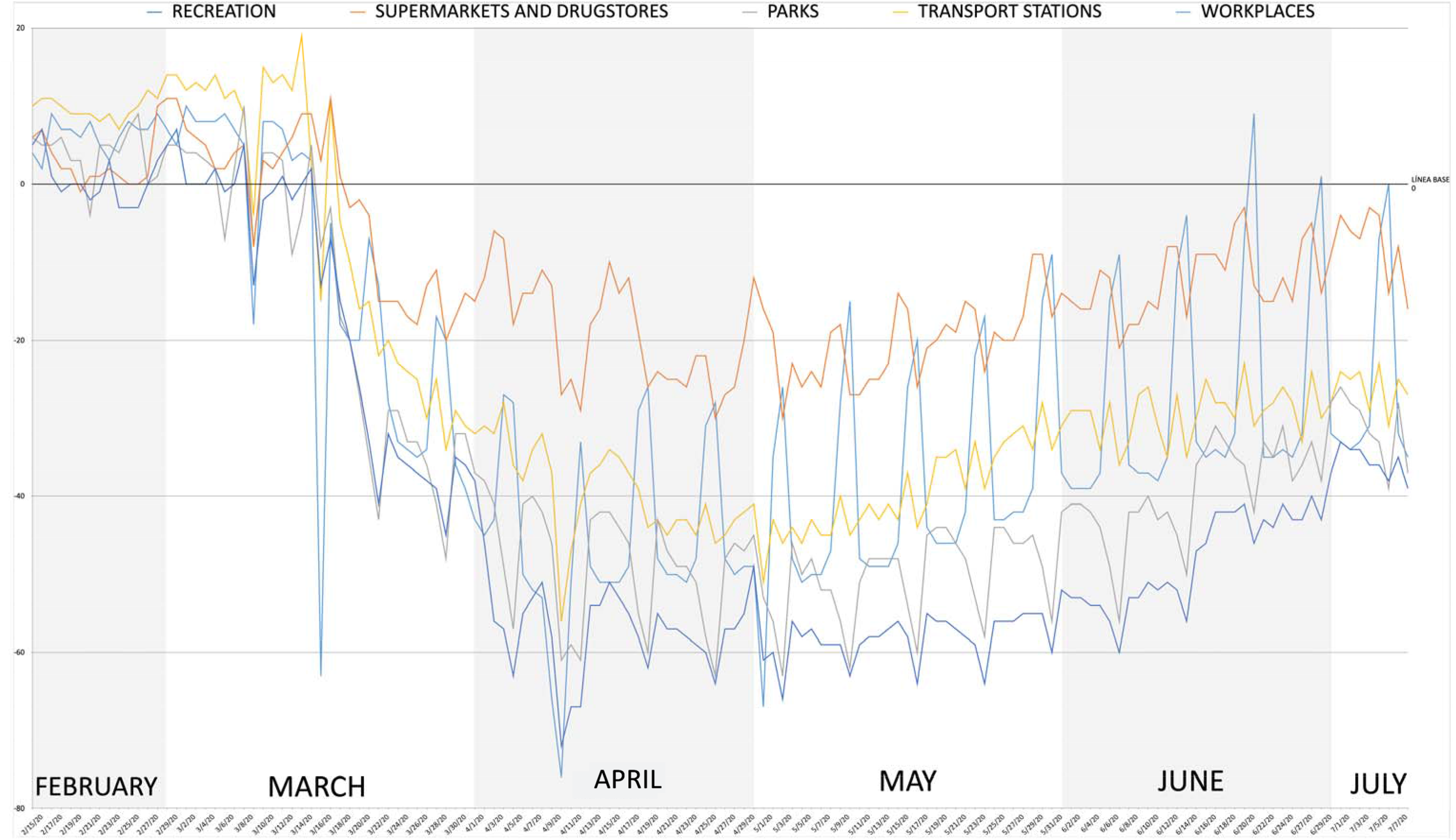
Mobility Levels using Google’s “Local Mobility Reports” Categorized

**Figure 3:**
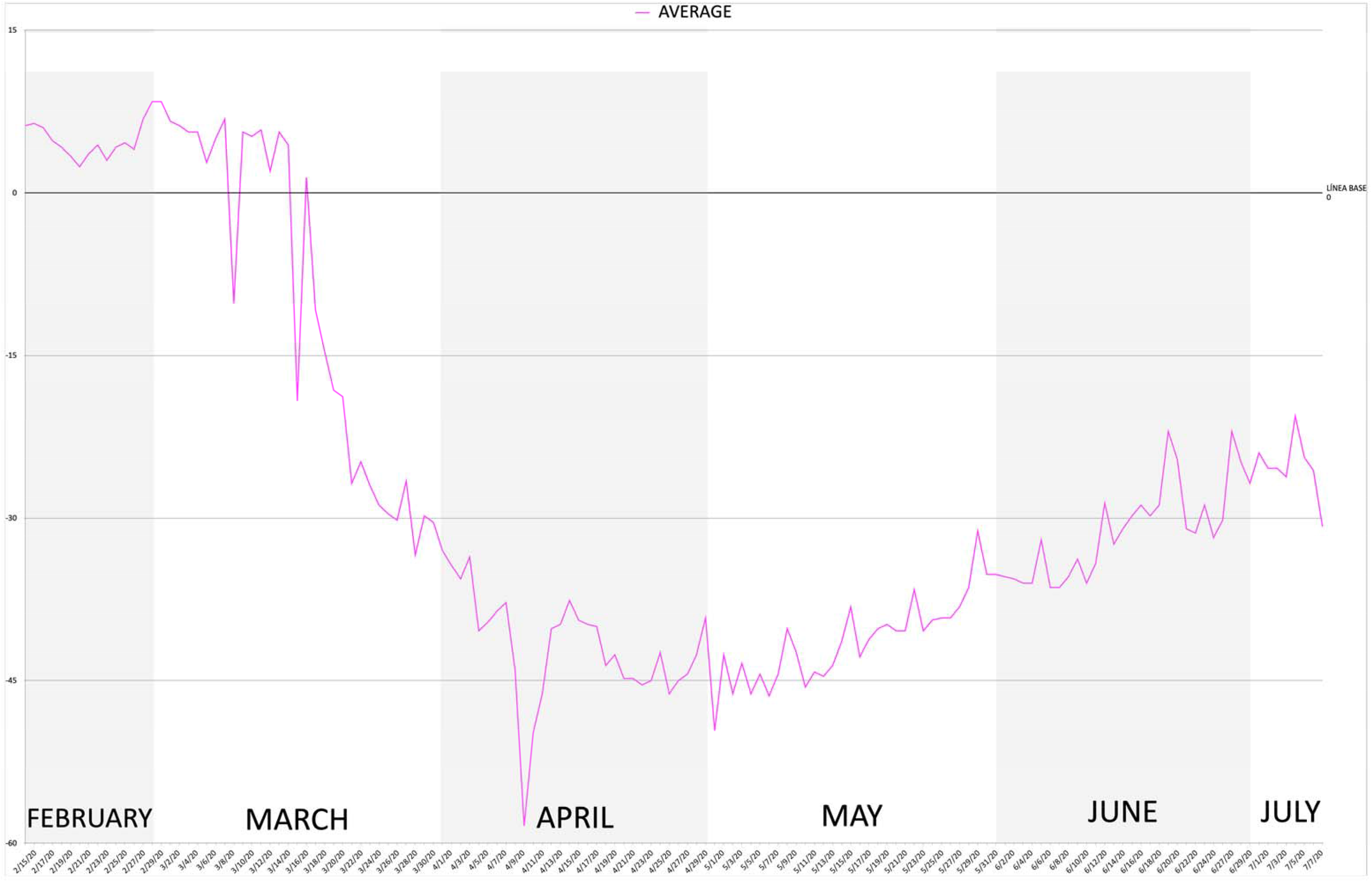
Average Mobility Levels using Google’s “Local Mobility Reports”

Figure 4 displays the reproductive numbers R0 and Re in Querétaro during the same time period of March 1 through July 5. We observe through the data a positive relationship between Re, R0, and Google mobility levels in Querétaro, Mx. This indicates that an increase in mobility in the population in Querétaro is associated with an increase in transmission of Covid-19. At the start of the lockdown, when social distancing measures were introduced the R0 and Re did not decrease greatly, as public health and government officials hoped in order to control the spread, however, it did not increase dramatically either. There is a large decrease in mobility levels, as displayed in Figure 2, from February that lasts until approximately May 1st, which corresponds to a R0 and Re of 1.17 to 1.87 (for both R0 and Re). While there is decreased mobility in Querétaro, it is clear that the R0 and Re of Covid-19 doesn t drastically increase. That being said however, after May 1 there is a steady increase in mobility levels. When looking at the effective reproductive numbers and the basic reproductive index, both Re and R0 begin to increase starting at about May 16. This is understandable, as it reflects the delay between infection and Covid-19 diagnosis, and then the lag between the testing and the official reporting of test results. The R0 and Re increase from 1.45 in May 16 to 3.81 and 3.59, respectively, on July 5 as mobility levels increased from −49.6 on May 1 to −20.6 on July 5. According to this data, easing mobility restrictions should be reconsidered until the R0 and Re can be reduced to below R = 1 so as to decrease the rapid transmission of Covid-19.

**Figure 4:**
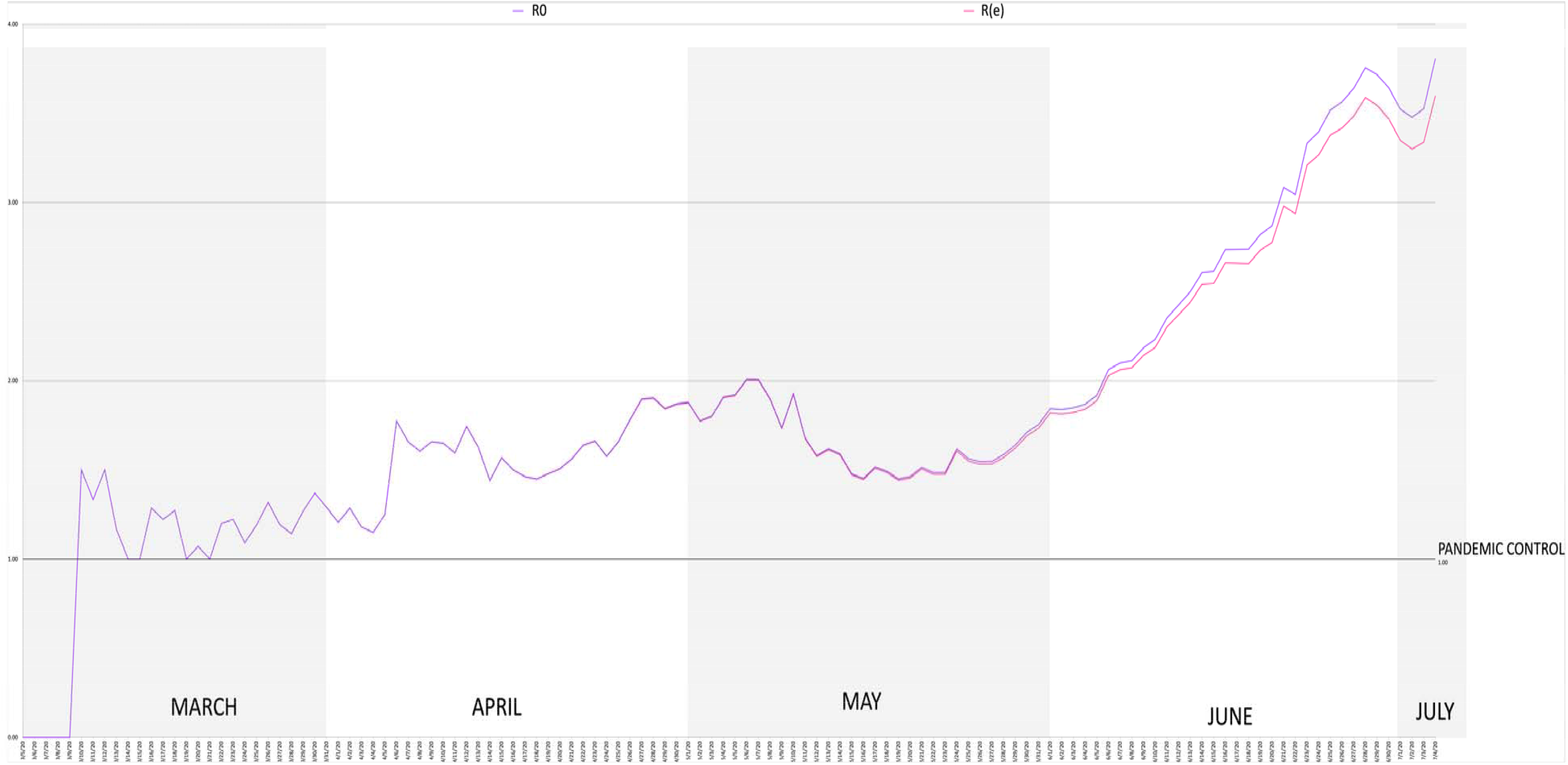
Reproductive Index & Effective Reproductive Numbers in Queretaro, Mexico

## Discussion of Mobility Levels & the Rate of Transmission of Covid-19 in Querétaro

So far, the interventions in place to contain the spread of Covid-19 have largely centered around non-pharmaceutical interventions, as there is no cure or vaccine, to decrease the number of contacts amongst individuals^3^. Some of these interventions include social distancing, wearing masks, prohibiting large public gatherings, and shutting down schools. In doing so, public health officials are attempting to halt modes of transmission amongst individuals in an effort to bring the R0 to below 1 to control and prevent the spread of SARS-CoV-2. This observational study attempts to describe and examine the relationship between mobility patterns in Querétaro, Mexico, and the rate of transmission of Covid-19. The observations show that when mobility levels were low in Querétaro, the reproductive index and effective reproductive number, R0 and Re, stayed relatively low—although never less than one. The non-pharmaceutical interventions have failed to reduce the R0 and Re in Queretaro. However, an increase in mobility levels that began in May brought a sharp increase in the rate of transmission, leading to an increase in Covid-19 cases. We predict that there will be an increase in the number of cases and deaths in Mexico if individual behavior at the community level isn’t taken. The estimated rate of transmission is calculated from the reported official numbers^16^. We acknowledge that the degree of underreporting Covid-19 deaths hasn’t been constant over time and could lead to an upwards bias of our R0 and Re estimates. Despite reductions and maintaining the reproductive number at a relatively stable number, it was insufficient in controlling the spread of Covid-19. Our observations suggest that in the absence of stringent measures that restrict mobility in Querétaro, the epidemic is bound to spread further. For instance, Lombardy, Italy, a region that was severely impacted by Covid-19 had a sharp decrease of 75% in the mobility of grocery stores and pharmacy, resulting in an estimated R0 of 0.58, therefore controlling the rate of transmission of the virus^19^. On the other hand, the lowest R0 in Querétaro, at the early stages of the epidemic, was at 1.00 and only for a few days. So, although the decrease in mobility levels in March and April helped to some extent stabilize the R0, it was not enough to control the spread. Only to exacerbated the rate of transmission once mobility levels increased again in May. Research suggests that individuals must change their behavior and limit mobility as best they can to decrease the reproductive index to less than one. We addressed how mobility levels are strongly correlated to the spread of transmission. But, for the R0 and Re to decrease to below 1, the population must holistically change their behavior (facemask, social distance, etc.) plus decreasing their mobility levels to help cut the rate of transmission and control the epidemic.

## Data Availability

The epidemiology data is public, gathered from the Federal Ministry of Health webpage (Direccion General de Epidemiologia. CONACYT, CentroGeo, GeoInt, DataLab. Gobierno Federal Mexico. Updated 15 June 2020. Gathered June 15th, 2020.
The mobility data, was gathered from Google's Local Mobility Reports for Queretaro, Mexico.  

https://coronavirus.gob.mx/datos/#DOView

https://www.google.com/covid19/mobility/

## Contributors and sources

The idea of writing about mobility and R0, along with the development of the pandemic in the community of Querétaro, Mexico arose after reading articles about this correlation in major cities in the United States. Dr. Isidro Guiterrez Alcocer gathered the team together and supervised the paper. Santiago Agraz gathered the data from the mobility trends presented by Google and from the official epidemiological data from the Mexican government s official COVID site, as well as using daily reports from the state government. Then, Oscar San Roman Orozco made the equations and graphs in order to obtain the R0 and Re. Vasiliki Radaios contribute to analysing the data, graphs, and formulating conclusions. All agreed on the final product and conclusions.

## Acknowledgements

We thank Dr. Teresa Garcia and Dr. Javier Avila, Dean and Academic Dean respectively of the Universidad Autonoma de Queretaro for all their support on the development of all projects related to the new COVID-19 Clinic of the University Health System.

To Dr. Juan Jose Moreno Ponce for his support and guidance.

## References

1. Bedford, J., Enria, D., Giesecke, J., Heymann, D. L., Ihekweazu, C., Kobinger, G.,… Technical Advisory Group for Infectious, H. (2020). COVID-19: towards controlling of a pandemic. Lancet; 395(10229), 1015–1018. doi: 10.1016/S0140-6736(20)30673-5

2. Wu, Z., & McGoogan, J. M. (2020). Characteristics of and Important Lessons From the Coronavirus Disease 2019 (COVID-19) Outbreak in China: Summary of a Report of 72314 Cases From the Chinese Center for Disease Control and Prevention. JAMA. doi: 10.1001/jama.2020.2648

3. A Wilder-Smith MD, DO Freedman, MD, Isolation, quarantine, social distancing and community containment: pivotal role for old-style public health measures in the novel coronavirus (2019-nCoV) outbreak, Journal of Travel Medicine, Volume 27, Issue 2, March 2020, taaa020, https://doi.org/10.1093/jtm/taaa020

4. Lauer SA, Grantz KH, Bi Q, et al. The Incubation Period of Coronavirus Disease 2019 (COVID-19) From Publicly Reported Confirmed Cases: Estimation and Application. Ann Intern Med 2020; 2019.

5. BBC, Staff. (February 29, 2020). Coronavirus en México: confirman los primeros casos de covid-19 en el país. Retrieved July 15, 2020. From. https://www.bbc.com/mundo/noticiasamerica-latina-51677751

7. Forbes Staff. (March 11, 2020). Querétaro confirma el primer caso de Covid-19. Retrieved June 10, 2020, from https://www.forbes.com.mx/actualidad-queretaro-confirma-casocovid-19/

8. World Health Organization. (2020). Coronavirus disease 2019 (COVID-19): situation report, 143.

9. Lai, A., Bergna, A., Acciarri, C., Galli, M., & Zehender, G. (2020). Early phylogenetic estimate of the effective reproduction number of SARS-CoV-2. Journal of medical virology, 92(6), 675–679.

10. Bergman, N., & Fishman, R. (2020). Correlations of Mobility and Covid-19 Transmission in Global Data. doi: 10.1101/2020.05.06.20093039.

11. Gandhi, R. T., Lynch, J. B., & del Rio, C. (2020). Mild or moderate COVID-19. New England Journal of Medicine.

12. Tolles J, Luong T. Modeling Epidemics With Compartmental Models. JAMA. Published online May 27, 2020. doi: 10.1001/jama.2020.8420.

13. Dirección Jurídica y Consultiva de la Secretaria de Gobierno del Poder Ejecutivo del Estado de Querétaro. Periódico Oficial La Sombra de Arteaga. “Acuerdo de medidas extraordinarias para mitigar la enfermedad COVID-19 y potencializar el distanciamiento social”. Published May 2, 2020. https://lasombradearteaga.segobqueretaro.gob.mx/04_period/frame.html

14. Kermack WO, McKendrick AG. A contribution to the mathematical theory of epidemics. Proc Royal Soc Math Phys Eng Sci. 1927;115(772):700–21.

15. Ridenhour, B., Kowalik, J. M., & Shay, D. K. (2018). El número reproductivo básico (R0): consideraciones para su aplicación en la salud póblica. American Journal of Public Health, 108(Suppl 6), S455–S465. https://doi.org/10.2105/AJPH.2013.301704s.

16. N. van Doremalen, T. Bushmaker, D.H. Morris, M.G. Holbrook, A. Gamble, B.N. Williamson, A. Tamin, J.L. Harcourt, N.J. Thornburg, S.I. Gerber, et al. Aerosol and surface stability of SARS-CoV-2 as compared with SARS-CoV-1. N Engl J Med (2020).

17. Dirección General de Epidemiología. CONACYT, CentroGeo, GeoInt, DataLab. Gobierno Federal México. Actualizada 15 Junio 2020. Consulted June 15th, 2020. https://coronavirus.gob.mx/datos/#DOView.

18. Pellis L, Ferguson NM, Fraser C. Threshold parameters for a model of epidemic spread among households and workplaces. J R Soc Interface. 2009;6 (40):979–87.

19. P Nouvellet, S Bhatia, A Cori et al. Reduction in mobility and COVID-19 transmission. Imperial College London 08–06-2020), doi: https://doi.org/10.25561/79643.

20. Miller, A. C., Foti, N. J., Lewnard, J. A., Jewell, N. P., Guestrin, C., & Fox, E. B. (2020). Mobility trends provide a leading indicator of changes in SARS-CoV-2 transmission. medRxiv, 2020.2005.2007.20094441. doi: 10.1101/2020.05.07.20094441

